# Correlation of COVID-19 Mortality with Clinical Parameters in an Urban and Suburban Nursing Home Population

**DOI:** 10.1101/2020.10.15.20213629

**Authors:** Richard S. Kirby, John A. Kirby

## Abstract

**Importance and Objective:** COVID-19 has a high mortality rate amongst nursing home populations (26.4% nationally and 28.3% in New Jersey). Identification of factors influencing mortality in COVID-19 positive nursing home populations may help direct physicians towards appropriate glycemic, blood pressure, weight, kidney function, lipid, thyroid, and hematologic management to reduce COVID-19 mortality.

**Design, Setting, and Participants:** Retrospective cross-sectional study of patients in two nursing home facilities (one urban, one suburban) from 3/16/2020 to 7/13/2020 with positive COVID-19 PCR assays. Age, race, sex, lipids, hematologic parameters, body mass index, blood pressure, thyroid function, albumin, blood urea nitrogen, creatinine, and hemoglobin A1c were correlated with COVID-19 mortality by chi-squared analysis.

**Main Outcome and Results:** 56 patients met the inclusion criteria for the study. Mortality was 14.3% while the New Jersey nursing home average mortality rate was 28.3% as of August 2020. Our patient cohort had a 49.5% reduction in mortality compared to the state average.

In our overall cohort, none of the clinical parameters correlated with COVID-19 mortality using chi-squared analysis. In the 56 patient cohort, average clinical and laboratory findings were 74.0 years, 62.5% female, 28.5% uncontrolled hypertension, BMI 25.6, hemoglobin A1c 6.4, TSH 2.4, vitamin B12 568.3, folate 12.4, iron 47.8, total iron binding capacity 271.8, hemoglobin 11.6, albumin 3.5, triglycerides 100.3, total cholesterol 133.5, HDL 40.9, and BUN to Creatinine ratio 22.2:1. Logistic multivariate regression analyses failed to demonstrate clinically significant correlation with COVID-19 mortality.

In the urban nursing home, BUN to creatinine ratio exceeding 20:1 was the only factor that showed statistical significance to COVID-19 mortality (p = 0.03). In the suburban nursing home, age over 80 was the only clinical factor demonstrating statistical significance to COVID-19 mortality (p = 0.003).

**Conclusions and Relevance:** In our COVID-19 positive nursing home patients, no one parameter was clinically significant in the overall 56-patient cohort; however, mortality in our population was 14.3% compared to New Jersey’s 28.3%, a 49.5% reduction in mortality. Rigorous control of the aforementioned clinical parameters may have contributed to this reduction in mortality. Further research requires analysis of more nursing home patients to determine whether rigorous control of clinical parameters decreases mortality from COVID-19.

**Key Points:** *Question:* What clinical parameters lead to a lower mortality rate in nursing home patients with COVID-19?

*Findings:* In this cross-sectional analysis of 56 SARS-CoV-2 positive New Jersey nursing home residents from March to July 2020, controlling hemoglobin A1c, blood pressure, hematologic and lipid panels to recommended levels yielded a mortality rate of 14.3%, a 49.5% reduction from the 28.3% mortality rate of COVID-19 in New Jersey nursing homes.

*Meaning:* Maintaining rigorous control of clinical parameters in nursing home populations may account for a decreased mortality rate of COVID-19.

## Introduction

COVID-19, is an emerging infectious disease with a particularly high mortality rate amongst nursing home populations compared to the general population (26.4% vs. 7.8% nationally and 28.3% vs. 7.6% in New Jersey as of August 5^th^, 2020).^1-4^ Nursing home populations consist of residents who have chronic underlying medical conditions such as hypertension, cardiac disease, pulmonary disease, diabetes mellitus, obesity, thyroid disease, hematologic disease, gastroesophageal reflux disease, and renal disease.^5^ Identifying clinical parameters that positively or negatively affect mortality from COVID-19 in a nursing home population should be of paramount importance to reducing the mortality rate for this vulnerable population.

Prior studies that discuss the prevalence of co-morbidities in the total COVID-19 positive population show that 60-90% of patients have a comorbidity.^6^ The co-morbidities for COVID-19 positive patients in the nursing home population include chronic lung disease (38%), diabetes (38%), cardiovascular disease (81%), cerebrovascular accident (40%), renal disease (38%), hemodialysis (6%), cognitive impairment (58%), and obesity (23%).^7^ These co-morbidity prevalence rates have only been reported in two case series so far.^7,8^ These case series fail to report the severity of those conditions and how that might have affected the overall mortality rate of the patients in those nursing homes.

Our study looks at the clinical parameters of COVID-19 positive patients in a skilled nursing facility and aims to identify significant factors that lead to increased or decreased mortality in the nursing home population. The purpose of the study is to direct nursing home physicians towards appropriate glycemic, blood pressure, weight, kidney function, lipid, thyroid, and hematologic management so that COVID-19 mortality can be reduced.

## Methods and Materials

### Subjects

Retrospective cross-sectional study of 56 nursing home residents from 3/16/2020 to 7/13/2020 who tested positive for COVID-19 via PCR assay. These patients were under the primary care of one internal medicine physician and lived in two nursing home facilities, one in an urban setting (sub-cohort comprised of 26 patients), and the other in a suburban setting (sub-cohort comprised of 30 patients) in New Jersey. Our study was approved by the Cooper University Hospital institutional review board. Written permission was granted by the nursing home administrators.

### Demographics

The median age of our patient cohort was 74.0 years with an interquartile range (IQR) of 64.8 to 83.0 years old. The patient cohort was 62.5% female, the median body mass index (BMI) was 25.6 with an IQR of 23.4 to 33.0, 61% of patients had diabetes, and 91% with hypertension defined as greater than 120/80 mm Hg.

### Procedures

A retrospective cross-sectional analysis. All patients (both symptomatic and asymptomatic) in both nursing facilities were tested for COVID-19 positivity during the time interval of our study. A list of patients with a positive COVID-19 PCR assay was obtained. From these patients, sex, age, blood pressure (BP), BMI, hemoglobin A1c, thyroid stimulating hormone (TSH), vitamin B12, folate, iron, total iron binding capacity (TIBC), hemoglobin, albumin, triglycerides, total cholesterol, low density lipoprotein (LDL), high density lipoprotein (HDL), blood urea nitrogen (BUN), and creatinine (Cr) were obtained from the patients’ hybrid PointClickCare electronic medical record and paper charts. BMI was calculated using height, weight, and a BMI calculator. Patients were treated using low molecular weight heparin therapy if not previously anticoagulated.^9^ Acetaminophen was provided on an as needed basis for fever.

### Definitions and Criteria

Triglycerides were labelled as normal if the value was between 35-160 mg/dL, and high if the value was over 160 mg/dL. Total cholesterol was labelled as normal if the value was 200 mg/dL or less, and high if the value was over 200 mg/dL. HDL cholesterol was normal if the value was between 35-80 mg/dL, low if the value was below 35 mg/dL, and high if the value was above 80 mg/dL. LDL cholesterol was normal if the value was at or below 130 mg/dL and high if the value was above 130 mg/dL. BMI was defined as normal if the value was between 20.0-24.9, overweight if the value was between 25.0-29.9, and obese if the value was above 30.0. Hypertension was defined as any blood pressure greater than 120 mm Hg systolic and 80 mm Hg diastolic. Uncontrolled hypertension was defined as any blood pressure greater than 130 mm Hg systolic and 90 mm Hg diastolic. Patients were considered non-diabetic if their hemoglobin A1c was less than 5.7%, pre-diabetic if their hemoglobin A1c was between 5.7%-6.4%, and diabetic if their hemoglobin A1c was above 6.4%. Albumin was normal if the value was between 3.5-5.5 mg/dL. BUN to Cr ratio was normal between 10-20:1 mg/dL. TSH was normal if the value was between 0.4-4.0 mU/L. Hemoglobin was normal if the value was between 12-17.5 g/dL. Vitamin B12 was normal if the value was between 200-900 pg/mL. Folate was normal if the value was between 2-20 ng/mL. Iron was normal if the value was between 50-170 mg/dL.

### Data Collection

All clinical parameter values were obtained from clinical and laboratory reports.

### Statistical Tests

Data was broken into categories as described above. COVID-19 mortality was analyzed against all other categories using chi-squared analysis. Logistic multivariate regression analyses were used to compare COVID-19 mortality, age, and all the other clinical parameters with a chi-squared p-value less than 0.30. A p-value less than or equal to 0.05 represents an acceptable level of statistical significance.

## Results

Three out of 26 patients died from COVID-19 in the urban nursing home setting and 5 out of 30 patients died from COVID-19 in the suburban nursing home location. Analysis of our whole patient cohort showed that the mortality of COVID-19 positive patients was 14.3% (8 out of 56 patients) compared to the 28.3% mortality rate observed throughout New Jersey nursing homes. Our 56 patient cohort had a 49.5% reduction in mortality compared to the state average.

Taking a closer look at our whole patient cohort, we analyzed age, sex, race, blood pressure, BMI, hemoglobin A1c, TSH, vitamin B12, folate, iron, TIBC, hemoglobin, albumin, triglycerides, total cholesterol, HDL, and BUN:Cr ratio and compared these parameters to COVID-19 mortality. Six/29 (20.7%) Black patients, 1/18 (5.6%) White patients, and 1/9 (11.1%) Hispanic patients died from COVID-19. Obesity was found in 28.6% of the patient cohort. Cardiovascular disease was present in 94.6% of the patient cohort. Cerebrovascular accident was present in 28.6% of the patient cohort. Hypertension was present in 85.7% of the patient cohort with 28.7% of the patients having uncontrolled hypertension defined as greater than 130/90 mm Hg. Hyperlipidemia was present in 67.9% of the patient cohort. Diabetes was present in 55.4% of the patient cohort. Chronic lung disease was present in 30.4% of the patient cohort. Renal disease was present in 42.9% of the patient cohort. We found that none of these factors had statistical significance towards increased COVID-19 mortality amongst this cohort (Table 1 and 2). Logistic regression analyses using combinations of the aforementioned parameters failed to demonstrate any statistically significant correlation with COVID-19 mortality.

**Table 1.** Combined Urban and Suburban Nursing Home Population Clinical Parameters Significance to COVID-19 Mortality using Chi-squared Analysis

| Clinical Parameter | Median/Prevalence | Average | p-value |
| --- | --- | --- | --- |
| Age (years) | 74.0 (IQR<sup>a</sup> = 64.8-83.0) | 73.8 | 0.055 |
| Race | 51.8% Black, 16.1% Hispanic, 32.1% White | 51.8% Black, 16.1% Hispanic, 32.1% White | 0.34 |
| Sex | 62.5% Female; 37.5% Male | 62.5% Female; 37.5% Male | 0.43 |
| Blood Pressure (mm Hg) | 125 SBP<sup>b</sup> (IQR<sup>a</sup> = 119-131); 72 DBP<sup>c</sup> (IQR<sup>a</sup> = 66-77) | 125 SBP<sup>b</sup>; 72 DBP<sup>c</sup> | 0.55 |
| BMI | 25.6 (IQR<sup>a</sup> = 23.4-33.0) | 28.4 | 0.51 |
| Hemoglobin A1c (%) | 6.4 (IQR<sup>a</sup> = 5.6-7.7) | 6.7 | 0.77 |
| TSH (mU/L) | 1.9 (IQR<sup>a</sup> = 1.4-2.5) | 2.4 | 0.26 |
| Vitamin B12 (pg/mL) | 506 (IQR<sup>a</sup> = 368.5-646.5) | 568.3 | 0.77 |
| Folate (ng/mL) | 13.4 (IQR<sup>a</sup> = 8.9-16.5) | 12.4 | 0.42 |
| Iron (mg/mL) | 47 (IQR<sup>a</sup> = 32.5-60.0) | 47.8 | 0.63 |
| TIBC (µg/dL) | 271 (IQR<sup>a</sup> = 234.0-310.3) | 271.8 | 0.92 |
| Hemoglobin (g/L) | 11.5 (IQR<sup>a</sup> = 10.5-12.8) | 11.6 | 0.97 |
| Albumin (mg/dL) | 3.5 (IQR<sup>a</sup> = 3.1-3.8) | 3.5 | 0.52 |
| Triglycerides (mg/dL) | 85 (IQR<sup>a</sup> = 61-141) | 100.3 | 0.35 |
| Total Cholesterol (mg/dL) | 129 (IQR<sup>a</sup> = 113-143) | 133.5 | 0.61 |
| HDL (mg/dL) | 41 (IQR<sup>a</sup> = 33-47) | 40.9 | 0.46 |
| LDL (mg/dL) | 72 (IQR<sup>a</sup> = 55.8-82.3) | 70.0 | 0.72 |
| BUN:Cr (mg/dL) | 20 (IQR<sup>a</sup> = 15.2-27.4) | 22.2 | 0.11 |

a = Interquartile Range
b = Systolic blood pressure
c = Diastolic blood pressure

**Table 2.** Clinical Parameter Comparison of COVID-19 Survival vs. Mortality in Combined Urban and Suburban Nursing Homes

| **Clinical Parameter** | **48 Survived COVID-19** | **8 Died from COVID-19** |
| --- | --- | --- |
| Age (years) | 72.1 | 83.8 |
| Race | 47.9% Black, 16.7% Hispanic, 35.4% White | 75% Black, 12.5% Hispanic, 12.5% White |
| Blood Pressure (mm Hg) | 125 SBP<sup>a</sup>, 72 DBP<sup>b</sup> | 124 SBP<sup>a</sup>, 71 DBP<sup>b</sup> |
| BMI | 29.2 | 23.6 |
| Hemoglobin A1c (%) | 6.8 | 6.2 |
| TSH (mU/L) | 2.6 | 1.7 |
| Vitamin B12 (pg/mL) | 571.4 | 545.2 |
| Folate (ng/mL) | 12.0 | 14.8 |
| Iron (mg/mL) | 48.1 | 46.2 |
| TIBC (µg/dL) | 272.6 | 267.8 |
| Hemoglobin (g/L) | 11.6 | 11.9 |
| Albumin (mg/dL) | 3.4 | 3.6 |
| Triglycerides (mg/dL) | 101.2 | 92.4 |
| Total Cholesterol (mg/dL) | 133.0 | 137.2 |
| HDL (mg/dL) | 40.5 | 43.8 |
| LDL (mg/dL) | 69.1 | 74.8 |
| BUN:Cr (mg/dL) | 22.2 | 22.2 |

In the urban nursing home facility and the suburban nursing home facility, clinical parameters mentioned above were compared to COVID-19 mortality via chi-squared analysis (Table 3). Only BUN to Cr ratio above 20:1 showed statistical significance to increased COVID-19 mortality (p = 0.03) in the urban nursing home facility and age over 80 (p = 0.003) showed significance to increased COVID-19 mortality in the suburban nursing home facility. Thirteen out of the 30 patients (43.3%) in the suburban nursing facility who contracted COVID-19 were over 80 years old. Five of these 13 patients (38.5%) died (all the patients who died in this facility). In the urban nursing home, 9 out of 26 patients (34.6%) who contracted COVID-19 were over 80 years old. None of these 9 patients died. Clinical parameters for patients over 80 years old were compared in Table 4. No statistical significance was found comparing clinical parameters and COVID-19 mortality for the greater than 80-year-old sub-cohort in the urban or suburban nursing home.

**Table 3.** Separate Urban and Suburban Nursing Home Population Clinical Parameters

| **Clinical Parameter** | **Urban Nursing Home Average (26 Patients)** | **Suburban Nursing Home Average (30 Patients)** |
| --- | --- | --- |
| Age (years) | 71.7 | 75.6* |
| Race | 61.5% Black, 15.4% Hispanic, 23.1% White | 43.3% Black, 16.7% Hispanic, 40.0% White |
| Blood Pressure (mm Hg) | 125 SBP<sup>a</sup>, 73 DBP<sup>b</sup> | 125 SBP<sup>a</sup>, 71 DBP<sup>b</sup> |
| BMI | 29.5 | 27.2 |
| Hemoglobin A1c (%) | 6.5 | 6.9 |
| TSH (mU/L) | 2.7 | 2.2 |
| Vitamin B12 (pg/mL) | 524.2 | 610.5 |
| Folate (ng/mL) | 11.2 | 13.1 |
| Iron (mg/mL) | 51.9 | 43.5 |
| TIBC (μg/dL) | 265.2 | 278.5 |
| Hemoglobin (g/L) | 11.2 | 12 |
| Albumin (mg/dL) | 3.5 | 3.5 |
| Triglycerides (mg/dL) | 94.8 | 106.0 |
| Total Cholesterol (mg/dL) | 130.2 | 137 |
| HDL (mg/dL) | 41.8 | 39.9 |
| LDL (mg/dL) | 67.4 | 72.3 |
| BUN:Cr (mg/dL) | 20.8* | 23.6 |

a = Systolic blood pressure
b = Diastolic blood pressure
\* = $p < 0.05$ determined by chi-squared analysis

**Table 4.** Separate Urban and Suburban Nursing Home Population Clinical Parameters for Patients over 80 Years Old

| **Clinical Parameter** | **Urban Nursing Home Average (9 Patients)** | **Suburban Nursing Home Average (13 Patients)** |
| --- | --- | --- |
| Race | 66.7% Black, 22.2% Hispanic, 11.1% White | 46.2% Black, 15.4% Hispanic, 38.5% White |
| Blood Pressure (mm Hg) | 126 SBP<sup>a</sup>, 74 DBP<sup>b</sup> | 124 SBP<sup>a</sup>, 69 DBP<sup>b</sup> |
| BMI | 28.1 | 26.9 |
| Hemoglobin A1c (%) | 5.7 | 6.5 |
| TSH (mU/L) | 2.8 | 2.1 |
| Vitamin B12 (pg/mL) | 547.6 | 728.6 |
| Folate (ng/mL) | 10.6 | 14.9 |
| Iron (mg/mL) | 52.9 | 37.5 |
| TIBC (µg/dL) | 288.4 | 265.9 |
| Hemoglobin (g/L) | 10.8 | 11.8 |
| Albumin (mg/dL) | 3.5 | 3.4 |
| Triglycerides (mg/dL) | 100.2 | 86.9 |
| Total Cholesterol (mg/dL) | 123.8 | 120.6 |
| HDL (mg/dL) | 46.3 | 40.6 |
| LDL (mg/dL) | 56.8 | 62.0 |
| BUN:Cr (mg/dL) | 25.9 | 24.9 |

a = Systolic blood pressure
b = Diastolic blood pressure

## Discussion

We found that in our whole patient cohort, the COVID-19 mortality rate was 14.3%, a 49.5% reduction in mortality compared to the 28.3% mortality rate from COVID-19 in the New Jersey nursing home population. Upon further investigation, there was no clinical or social parameter (race, sex, age, hypertension, BMI, hemoglobin A1c, TSH, vitamin B12, folate, iron, TIBC, hemoglobin, albumin, triglycerides, total cholesterol, HDL, and BUN to Cr ratio) in our entire cohort that indicated increased or decreased likelihood of COVID-19 mortality. In the urban nursing home facility, BUN to Cr ratio over 20:1 was the only clinical parameter that indicated increased likelihood of COVID-19 mortality (p = 0.03). In the suburban nursing home facility, age over 80 years was the only clinical parameter that showed increased likelihood of COVID-19 mortality (p = 0.003) because all the patients who died in this facility were over the age of 80. No specific clinical parameter in the over 80-year-old suburban nursing home sub-population increased mortality.

This dramatically lower mortality for our combined group of nursing home patients may provide guidance in reducing COVID-19 mortality without the discovery of new treatments. The nursing home residents from our cohort on average had rigorous control of their diabetes, blood pressure, weight, lipids, and hemoglobin. The cohort’s median hemoglobin A1c for diabetics was 6.4% which is excellent control according to the American Diabetes Association which recommends controlling hemoglobin A1c to less than 8.0% for older patients with multiple co-morbidities.^10^ The cohort’s average blood pressure was 125 mm Hg systolic BP and 72 mm Hg diastolic BP. According to the Joint National Commission 8 guidelines on hypertension which recommends that the general population over age 60 have blood pressure less than 150 for systolic BP and 90 for diastolic BP, the cohort had excellent BP control at 125 systolic BP and 72 diastolic BP.^11^ The cohort’s median BMI was 25.6 which was fair control according to the Systematic Evidence Review from the Obesity Expert Panel which recommends that adults maintain a BMI between 18.5 and 25.0.^12^ LDL cholesterol average was 70.0 which was controlled to below the upper limit of normal for the 67.9% of the patient cohort with hyperlipidemia. All other clinical parameters were controlled to their normal reference ranges except for hemoglobin in which the average was 11.6 g/L which is under the lower limit of normal at 12.5 g/L.

Nursing home patient clinical parameter control and its effects on COVID-19 have not been explored in the literature so far. A few papers detailed the rates of underlying conditions in a COVID-19 positive nursing home population.^7,8^ Cardiovascular disease, cerebrovascular accident, chronic lung disease, diabetes, renal disease, and obesity had prevalence rates of 81%, 40%, 38%, 38%, 38%, and 23% respectively in the COVID-19 positive nursing home population.^7^ These are similar rates seen in our patient cohort: 94.6%, 28.6%, 30.4%, 55.4%, 42.9%, and 28.6% for cardiovascular disease, cerebrovascular accident, chronic lung disease, diabetes, renal disease, and obesity respectively. Therefore, we can assume that our nursing home population has a co-morbidity rate similar to that of other nursing homes in the United States.

The 49.5% reduction of mortality in our patient population compared to the state average suggests that meticulous control of clinical parameters might help to decrease COVID-19 mortality in the nursing home population, regardless of race or sex. While we could not identify specific parameters in our combined urban and suburban populations that indicate higher risk of COVID-19 mortality, we believe it is imperative that physicians maintain strict control of their nursing home patients’ clinical parameters in order to mitigate the potential lethality of COVID-19.

### Study strengths and limitations

All patients in the cohort were treated under the auspices of a single physician, who provided uniform care to all COVID-19 positive patients. Data was collected from two nursing home facilities. A large-scale review of nursing home charts from multiple physicians in multiple nursing homes across New Jersey (and perhaps nationwide) is needed for comparison between our cohort’s clinical parameters and those of other physicians in other nursing homes in New Jersey and nationally. However, despite this limited data set, we feel that the finding of decreased mortality in our cohort compared to New Jersey’s COVID-19 positive nursing home population is supported by the rigorous clinical parameter control detailed in Table 1.

## Conclusion

From our data, we conclude that the 49.5% reduction of mortality in our nursing home cohort compared to the state average was a result of carefully controlled blood pressure, diabetes, lipids, and other clinical parameters. Future directions should include a large-scale review of nursing home patient clinical parameters to determine the specific factors in the nursing home population that increase COVID-19 mortality.

## Data Availability

All data was de-identified and is available upon request from the authors

## Abbreviations used in this paper

BMI: Body Mass Index
BP: Blood Pressure
BUN: Blood Urea Nitrogen
COVID-19: Coronavirus Disease 2019
Cr: Creatinine
HDL: High-density lipoprotein
LDL: Low-density lipoprotein
SARS-CoV-2: Severe Acute Respiratory Syndrome Coronavirus 2
TIBC: Total iron binding capacity
TSH: Thyroid Stimulating Hormone.

## Notes

### Competing Interest Statement

The authors have declared no competing interest.

### Funding Statement

This work was not supported with grants.

### Author Declarations

Cooper University Hospital IRB

## References

1. New Jersey Department of Public Health. New Jersey COVID-19 Cases Associated with LTC Outbreaks (As of 8/5/2020). https://www.state.nj.us/health/healthfacilities/documents/LTC_Facilities_Outbreaks_List.pdf. Accessed August 6, 2020.

2. Centers for Disease Control and Prevention. COVID-19 Nursing Home Dataset – Archived Data – Week Ending 8/2/20. https://data.cms.gov/Special-Programs-Initiatives-COVID-19-Nursing-Home/COVID-19-Nursing-Home-Dataset-Archived-Data-Week-E/i47f-65rd. Accessed August 6, 2020.

3. Centers for Disease Control and Prevention. COVIDView Summary ending on August 1, 2020. https://www.cdc.gov/coronavirus/2019-ncov/covid-data/covidview/past-reports/08072020.html. Accessed August 6, 2020.

4. New Jersey Department of Public Health. New Jersey COVID-19 Dashboard. https://www.nj.gov/health/cd/topics/covid2019_dashboard.shtml. Accessed August 6, 2020.

5. Moore KL, Boscardin WJ, Steinman MA, Schwartz JB. Patterns of chronic co-morbid medical conditions in older residents of U.S. nursing homes: differences between the sexes and across the agespan. J Nutr Health Aging. 2014;18(4):429–436.

6. Wiersinga WJ, Rhodes A, Cheng AC, Peacock SJ, Prescott HC. Pathophysiology, Transmission, Diagnosis, and Treatment of Coronavirus Disease 2019 (COVID-19): A Review. JAMA. 2020;324(8):782–793.

7. Arons MM, Hatfield KM, Reddy SC, et al. Presymptomatic SARS-CoV-2 Infections and Transmission in a Skilled Nursing Facility. N Engl J Med. 2020;382(22):2081–2090.

8. McMichael TM, Currie DW, Clark S, et al. Epidemiology of Covid-19 in a Long-Term Care Facility in King County, Washington. N Engl J Med. 2020;382(21):2005–2011.

9. Brouns SH, Brüggemann R, Linkens AEMJH, et al. Mortality and the Use of Antithrombotic Therapies Among Nursing Home Residents with COVID-19. J Am Geriatr Soc. 2020;68(8):1647–1652.

10. American Diabetes Association. 12. Older Adults: Standards of Medical Care in Diabetes-2020. Diabetes Care. 2020;43(Suppl 1):S152–S162.

11. James PA, Oparil S, Carter BL, et al. 2014 Evidence-Based Guideline for the Management of High Blood Pressure in Adults: Report From the Panel Members Appointed to the Eighth Joint National Committee (JNC 8). JAMA. 2014;311(5):507–520.

12. National Heart, Lung, and Blood Institute. Managing Overweight and Obesity in Adults: Systematic Evidence Review From the Obesity Expert Panel, 2013. http://www.nhlbi.nih.gov/guidelines/obesity/ser/index.htm. Accessed August 6, 2020.

